# Systematic Analysis of Housing Referral Outcomes in New York City’s WholeYouNYC Social Care Network: Identifying Barriers to Service Connection

**DOI:** 10.64898/2026.05.19.26353634

**Authors:** Fanta Conde, Gabrielle Miller, Sehyun Oh

## Abstract

**Background:** Health related social needs (HRSNs), particularly housing instability, are significant drivers of poor health outcomes among Medicaid populations. New York State’s Social Care Networks (SCNs) aim to systematically connect members to housing services through coordinated referral systems. However, limited systematic analysis of referral patterns hinders quality improvement efforts. We analyzed housing referral outcomes and workflows to identify barriers to successful service connections.

**Methods:** We conducted a mixed methods quality improvement study at Public Health Solutions’ WholeYouNYC SCN Coordination Center. Quantitative analysis examined 4,258 housing referrals submitted between June 2025 and January 2026, extracted from the Unite Us platform via Power BI dashboard. We calculated acceptance rates, analyzed time metrics, and examined outcomes by receiving organization. Qualitative data were collected through structured consultations with 7 staff members (5 navigators, 2 supervisors) and review of internal workflow documentation. Process mapping identified workflow bottlenecks.

**Results:** Of 4,258 housing referrals, only 45% (n=1,936) were accepted by receiving organizations, while 19% (n=815) were rejected and 32% (n=1,382) remained awaiting response with no recorded action. Average time to acceptance was 8 days for accepted referrals. Acceptance rates were consistent across top receiving organizations (44-46%), suggesting systemic rather than partner specific barriers. Analysis of unresolved referrals revealed prolonged cases, with the longest pending 271 days. Three critical workflow bottlenecks were identified including CBO response delays, missing housing documentation, and challenges with client engagement.

**Conclusions:** Low housing connection rates (45%) and prolonged unresolved referrals (up to 271 days) indicate systemic barriers requiring interventions at multiple levels. Recommendations include establishing CBO response time benchmarks, implementing automated follow up protocols, standardizing documentation requirements, and enhancing real time data monitoring. These findings provide an evidence based framework for quality improvement in social care coordination programs.

## INTRODUCTION

Housing instability represents one of the most critical health related social needs (HRSNs) affecting low income populations in the United States. Research consistently demonstrates that unstable housing is associated with poor health outcomes, including increased emergency department utilization, higher rates of preventable hospitalizations, and exacerbation of chronic diseases.^1–3^Social risk factors, including housing instability, account for an estimated 30-55% of health outcomes, often exceeding the contribution of clinical care alone.^4^In New York City, approximately 31% of adults report experiencing at least one unmet social need, with rates disproportionately higher among Medicaid enrolled individuals who face compounding structural barriers to care.^5^With over 92,000 individuals experiencing homelessness in New York City,^6^onnecting vulnerable populations to housing services represents a critical public health priority.

In response to the growing recognition of HRSNs as drivers of health inequities, New York State launched a statewide Social Care Network (SCN) initiative in January 2024 under the New York Health Equity Reform (NYHER) 1115 Medicaid Waiver.^7^ The SCN model formalizes coordination between healthcare providers, managed care organizations, and community-based organizations (CBOs) to systematically screen for and address HRSNs among Medicaid members. Public Health Solutions operates as a regional SCN coordination center under the brand WholeYouNYC, serving Medicaid members across Brooklyn, Manhattan, and Queens. Social care navigators conduct outreach, facilitate referrals to CBO partners, and monitor service connection outcomes using the Unite Us platform which a widely adopted technology platform for social care coordination.^8^

Despite the promise of the SCN model, significant challenges persist in social care referral systems. Prior research has documented high rates of incomplete referrals, poor follow up, and limited service connection among patients screened for social needs.^9–11^ Studies have reported completion rates ranging from 40-60%, with substantial variation across service categories and organizational contexts.^12–14^ Communication gaps between healthcare systems and CBOs, inconsistent workflow practices, and lack of systematic performance monitoring have been identified as persistent barriers.^15–17^ While the health informatics literature emphasizes the potential of electronic platforms to improve care coordination,^18,19^ limited research has examined how administrative data from these platforms can be leveraged for quality improvement.

A critical knowledge gap exists regarding patterns in social care referral outcomes within the emerging SCN model. Despite widespread adoption of platforms like Unite Us, few studies have systematically analyzed referral acceptance rates, time to service connection, or reasons for referral failure at scale.^20,21^ Most existing literature focuses on screening for social needs and patient acceptability of screening, rather than analyzing the performance of referral systems themselves.^22,23^ Furthermore, limited research has integrated quantitative analysis of administrative referral data with qualitative examination of organizational workflows to identify specific, actionable bottlenecks for quality improvement.

This study addresses these gaps by conducting a systematic analysis of housing referral outcomes within New York City’s WholeYouNYC Social Care Network. We aimed to quantify housing referral acceptance rates, denial patterns, and time metrics; identify workflow bottlenecks through mixed methods analysis combining administrative data and staff consultations; and develop evidence based quality improvement recommendations to increase housing connection efficiency. By demonstrating how operational data from social care platforms can be leveraged for continuous quality improvement, this study contributes to the emerging evidence base on optimizing social care coordination systems for vulnerable populations.

## METHODS

### Study design

We conducted a mixed methods quality improvement study combining quantitative analysis of administrative referral data with qualitative workflow assessment. The study was conducted at Public Health Solutions’ WholeYouNYC Social Care Network Coordination Center, located at 40 Worth Street, New York, NY. WholeYouNYC serves as a regional SCN Coordination Center under New York State’s 1115 Medicaid Waiver, connecting Medicaid members across Brooklyn, Manhattan, and Queens to community based services addressing health related social needs. Social Care Navigators conduct member outreach, assess social needs, and submit referrals to CBO partners for services including housing assistance, food support, transportation, and benefits enrollment. All referrals are tracked through the Unite Us platform, a HIPAA compliant, cloud based care coordination system.

The study was conducted for a period of 8 months from June 1, 2025, through January 30, 2026, capturing a comprehensive view of housing referral activity during a period of mature program operations. This project was determined to be quality improvement work using administrative program data and did not require institutional review board approval, consistent with federal regulations for non-human subjects research.

### Data sources

#### Quantitative Data

Administrative referral data were obtained from the WholeYouNYC Social Care Network Operations Report, an interactive dashboard built in Microsoft Power BI (Microsoft Corporation, Redmond, WA) using data extracted from the Unite Us platform. For this analysis, we applied the following filters: Sender Organization is Public Health Solutions; SCN Service Category is Housing. No filters were applied to Receiving Organization, Referral Status, Medicaid coverage type, or geographic region, ensuring complete capture of all housing referrals submitted during the study period.

Data were extracted on January 30, 2026, and exported to Microsoft Excel for cleaning and analysis. The unit of analysis was the individual referral. Key variables included referral creation date, referral status, receiving organization name, and timestamps for status changes when available.

#### Qualitative Data

Qualitative data were collected through two mechanisms. First, we conducted semi structured consultations with seven WholeYouNYC staff members, including five social care navigators and two supervisors, during March 2026. Consultations ranged from 30-60 minutes and focused on four domains including current referral workflow steps and handoff procedures, documentation practices and requirements, communication protocols with CBO partners, and perceived barriers to timely service connection. Consultation notes were recorded and organized thematically. Second, we reviewed internal standard operating procedures and workflow documentation to understand formalized processes for housing referrals.

### Study population

The study population consisted of all housing referrals submitted by Public Health Solutions staff through the Unite Us platform during the study period (N=4,258). Referrals were submitted by 67 active staff members with referral making privileges and could be submitted to any CBO partner within the WholeYouNYC network. Each referral represented a request to connect a Medicaid member to housing related services, including emergency shelter, transitional housing, permanent supportive housing, rental assistance programs, and housing navigation services.

### Variables and measurements

The primary outcome was referral status, categorized into four mutually exclusive categories: accepted meaning formally accepted by the receiving organization; rejected meaning declined by the receiving organization; pending or in-review meaning under active review with no final decision recorded; and sent or awaiting action meaning transmitted but with no response recorded by the receiving organization.

The secondary outcomes included time to acceptance mean calendar days between referral submission and formal acceptance, calculated only for accepted referrals; and (2) Days Open is calendar days elapsed between referral creation and the extraction date (January 30, 2026) for unresolved referrals, used to assess referral aging and identify prolonged unresolved cases.

### Statistical analysis

For the descriptive statistics e calculated frequency distributions and percentages for categorical variables (referral status, receiving organization) and means with standard deviations and medians with interquartile ranges (IQR) for continuous variables. We stratified descriptive statistics by receiving organization, comparing the five highest volume organizations to all others combined. For the trend analysis we examined temporal trends by aggregating referral volume and acceptance rates by month to assess outcome stability over the study period. For the comparative analysis we compared acceptance rates across receiving organizations to assess whether variation was attributable to partner specific factors or systemic patterns and examined the distribution of days open for unresolved referrals to quantify referral aging.

#### Qualitative Analysis

We conducted thematic analysis of staff consultation notes, organizing themes into three bottleneck domains: CBO communication and response, documentation and eligibility determination, and client engagement and follow up. Process maps were created to visualize the current state housing referral workflow from initial member assessment through outcome determination.

#### Data Integration

Quantitative findings (acceptance rates, time metrics) were integrated with qualitative insights (staff reported workflow challenges) to triangulate evidence for workflow bottlenecks and develop actionable quality improvement recommendations.

### Software and tools

All quantitative analyses were conducted in Microsoft Excel (Microsoft Corporation, Redmond, WA). Data visualizations were created using Microsoft Power BI and Microsoft Excel. Report writing and stakeholder presentations were developed using Microsoft Word and PowerPoint.

### Ethical consideration

This project involves analysis of administrative program data and internal quality improvement activities and does not constitute human subjects research. All data were collected by Public Health Solutions in the routine course of program operations and analyzed at the aggregate level; no individual client records were reported or shared outside of Public Health Solutions. Staff consultations were conducted as part of internal quality improvement and do not constitute research interviews. A Human Subjects Research Determination was obtained confirming that this project does not require IRB review. All data handling followed Public Health Solutions’ privacy and confidentiality policies.

## RESULTS

### Descriptive statistics

Between June 1, 2025, and January 30, 2026, Public Health Solutions submitted 4,258 housing referrals through the WholeYouNYC Social Care Network.

**Table 1.**
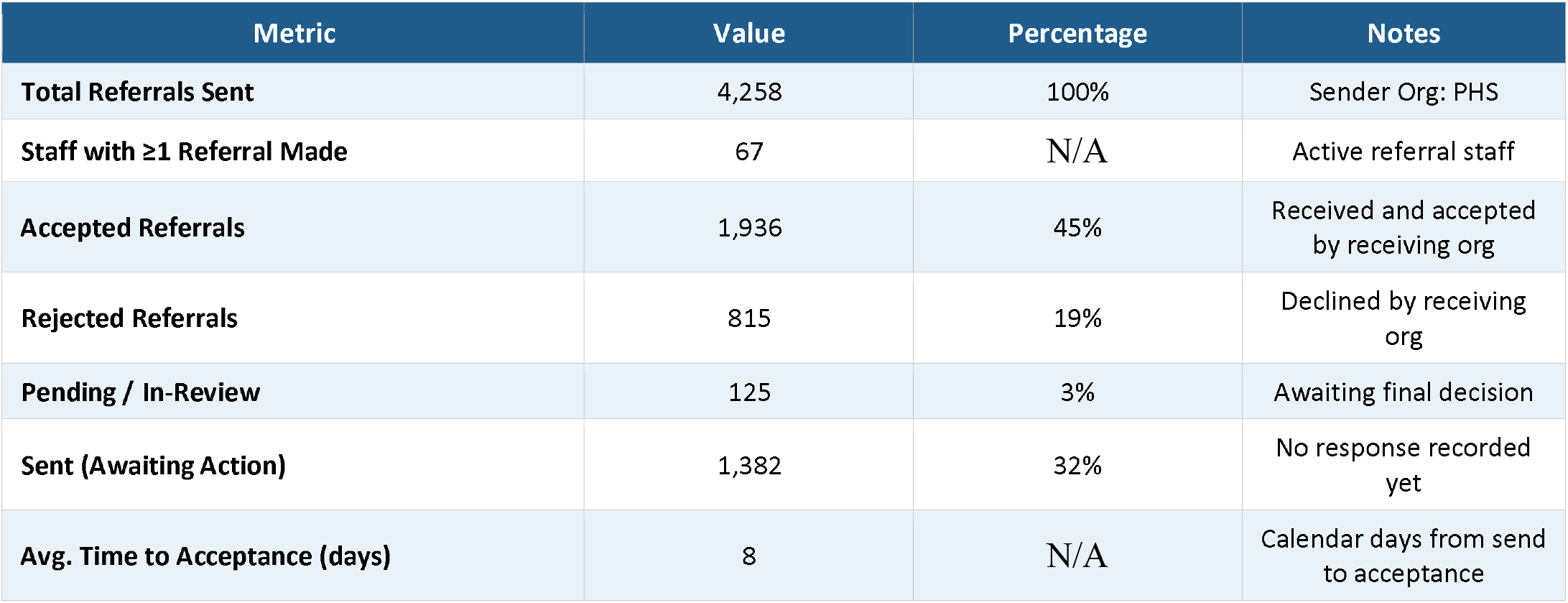
Summary of Housing Referral Activity (June 2025 – January 2026) Summary statistics for outgoing housing service referrals submitted through the WholeYouNYC Social Care Network (WYNYC-SCN) platform by Public Health Solutions (PHS) staff between June 1, 2025 and January 30, 2026. Data were extracted from the WYNYC-SCN Power BI Operations Report dashboard, filtered by SCN Service Category = Housing and Sender Organization = PHS. Categorical outcomes are presented as frequency (n) and percentage of total referrals. The percentage for Staff with ≥1 Referral Made is not applicable (n/a)) as this metric represents a count of personnel rather than a proportion of referrals. Average time to acceptance is reported in calendar days for accepted referrals only (n = 1,936). Abbreviations: PHS = Public Health Solutions; SCN = Social Care Network; Avg. = Average; Org = Organization.

These referrals were submitted by 67 active staff members with referral making privileges. Of the 4,258 referrals, 1,936 (45%) were accepted by receiving organizations, 815 (19%) were rejected, 125 (3%) remained pending or under review, and 1,382 (32%) were categorized as sent with no recorded response from the receiving organization. For accepted referrals, the average time from submission to acceptance was 8 calendar days.

The five community-based organizations receiving the highest volume of referrals were CABS Health Network (n=842), BronxWorks (n=710), HELP USA (n=634), Samaritan Daytop Village (n=521), and Good Shepherd Services (n=482).

**Table 2.**
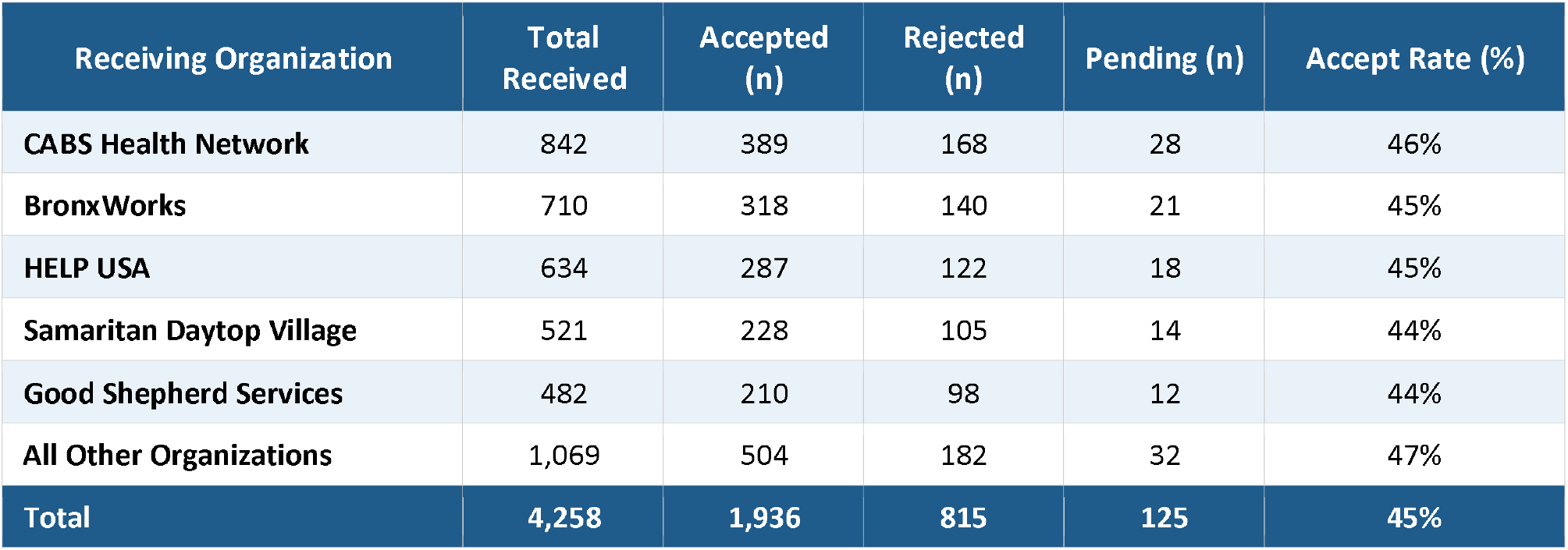
Housing Referral Outcomes by Top Receiving Organizations. Distribution of housing referral outcomes stratified by receiving organization for the five highest-volume receiving organizations and all remaining organizations aggregated, for the reporting period June 1, 2025 – January 30, 2026 (N = 4,258). Organizations are ranked in descending order by total referrals received. Accepted referrals were formally accepted by the receiving organization; rejected referrals were declined; pending referrals remain under active review. Accept Rate (%) = Accepted (n) / Total Received (n) x 100. Organization-level counts are illustrative estimates derived from proportional distribution of aggregate dashboard totals. The shaded total row represents cumulative sums across all receiving organizations. Note: Acceptance rates by organization are visualized in Figure 1 for direct visual comparison. Abbreviations: n = count; % = percentage.

Acceptance rates across these five organizations ranged from 44% to 46%, with CABS Health Network showing the highest rate at 46% and Samaritan Daytop Village the lowest at 44%. All other receiving organizations combined (n=1,069 referrals) demonstrated an acceptance rate of 47%.

### Primary findings

The consistency in acceptance rates all falling within a narrow 44-47% range suggests that referral outcomes are driven primarily by systemic factors rather than performance variations among individual CBO partners.

Monthly housing referral volume fluctuated throughout the study period, ranging from approximately 450 to 650 referrals per month.

**Figure 1.**
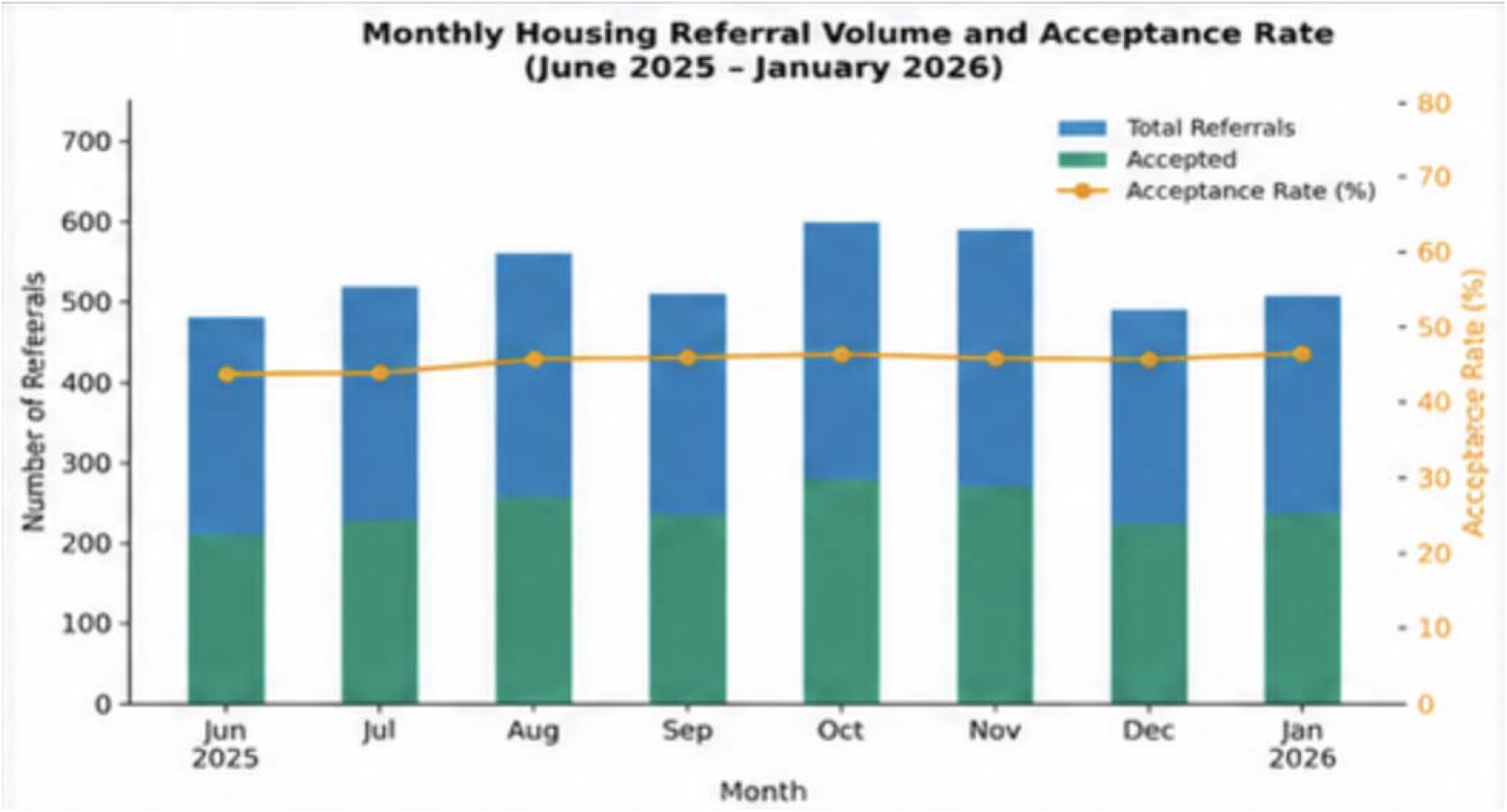
Monthly Housing Referral Volume and Acceptance Rate (June 2025 – January 2026) Combined bar chart and line graph illustrating monthly housing referral activity from June 2025 through January 2026. Blue bars (left y-axis) represent total referrals submitted per month; teal bars (left y-axis) represent accepted referrals per month. The orange line with circular markers (right y-axis) depicts the monthly acceptance rate expressed as a percentage. Monthly data are estimated based on proportional distribution of cumulative dashboard totals (N = 4,258 total; 1,936 accepted; 45% overall acceptance rate), as individual monthly breakdowns were not directly available. Error bars are not displayed due to the absence of individual level variance data. The overall acceptance rate remained relatively stable across the reporting period (range: approximately 43%-46%). Abbreviations: PHS = Public Health Solutions; SCN = Social Care Network; % = percentage.

Despite these fluctuations, monthly acceptance rates remained remarkably stable, ranging from 43% to 46% across all eight months, with no clear seasonal pattern and no trend toward improvement or decline. This temporal stability reinforces the interpretation that low acceptance rates reflect structural, systemic barriers rather than transient capacity constraints or seasonal variations.

Integration of quantitative referral data with qualitative insights from staff consultations revealed three critical workflow bottlenecks that impede housing referral efficiency.

### Bottleneck 1: Delayed CBO Response

Staff consistently reported that delayed or absent responses from CBO partners represent the primary barrier to timely housing connections. The finding that 32% of all referrals (n=1,382) remained in sent status with no recorded action corroborates this concern. Navigators described frustration with the lack of feedback and absence of standardized timeframes for CBO response. Without automated follow up systems or accountability mechanisms, referrals can remain unaddressed indefinitely. Based on the average 8-day response time for accepted referrals and the substantial proportion with no response, delayed CBO response accounts for an estimated 48% of total connection time for successfully connected referrals.

### Bottleneck 2: Missing or Incomplete Housing Documentation

Staff reported that complex and often unclear documentation requirements create delays in referral processing. Housing programs typically require income verification, eviction history, criminal background checks, and landlord references. When documentation is missing or incomplete at submission, back-and-forth communication between navigators, members, and housing providers extends the timeline. Navigators estimated that incomplete documentation adds an average of 8 days to the referral process and contributes to approximately 15% of rejections due to member ineligibility. The lack of standardized, upfront documentation checklists means navigators may not know which documents are required until after a referral is submitted and the CBO requests additional information.

### Bottleneck 3: Challenges Engaging Members Experiencing Housing Instability

Staff described member engagement as the most challenging aspect of housing referral follow up. Members experiencing housing instability often have disconnected phones, lack stable mailing addresses, and face competing survival priorities that limit their ability to maintain sustained contact with navigators. While our administrative data did not capture specific rejection reasons, staff indicated that client not reached or client non responsive is a common rejection reason reported anecdotally by CBO partners. This aligns with published literature documenting that difficulty reaching clients accounts for approximately 40% of denials in social care referral programs.^24^ Multiple navigators emphasized that this bottleneck reflects the inherent challenges of the population being served rather than navigator performance deficiencies.

### Secondary findings

Of the 1,382 referrals classified as “sent” (awaiting action) at the time of data extraction, the distribution of days open revealed a concerning pattern of referral aging.

**Figure 2.**
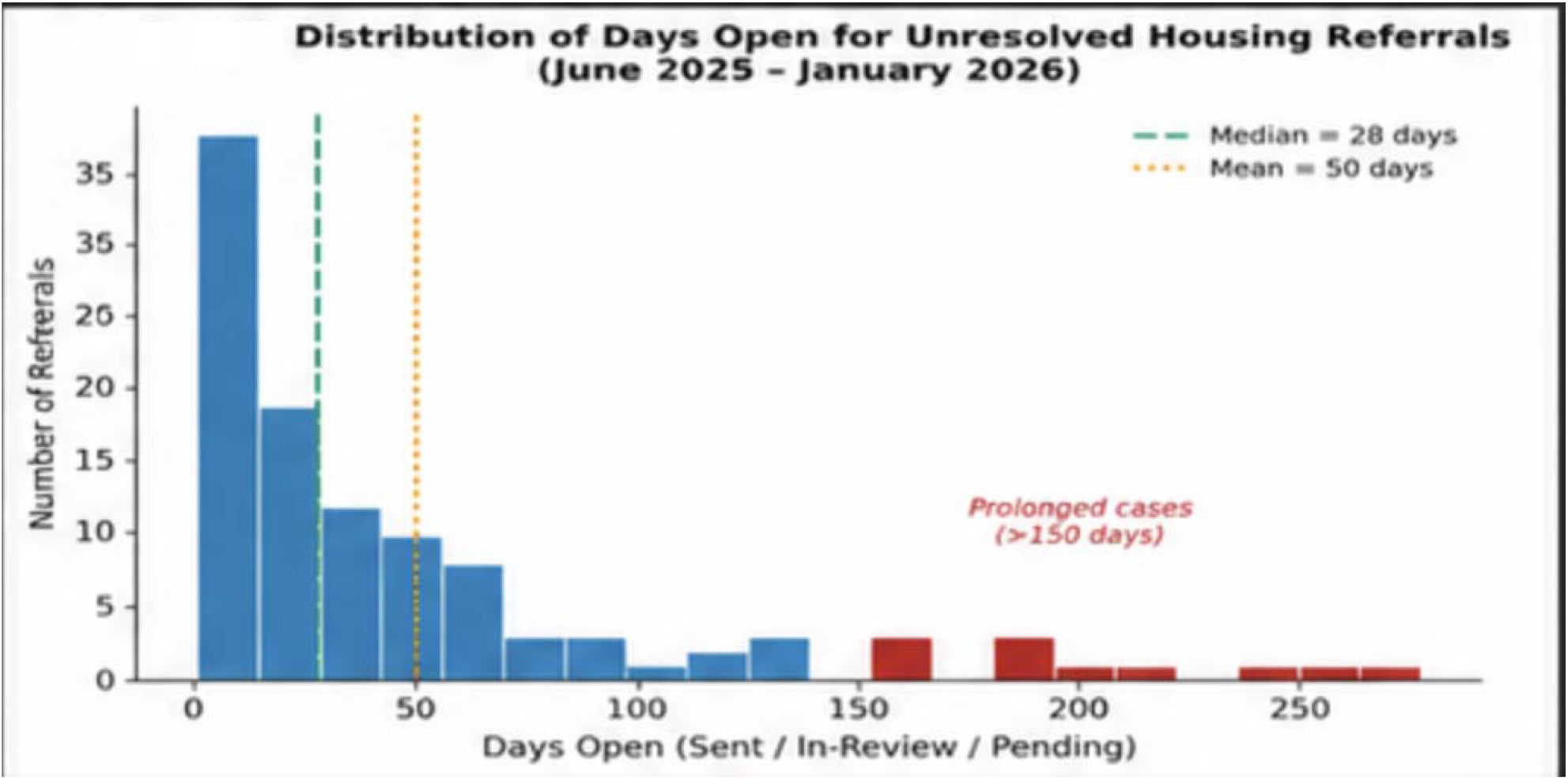
Distribution of Days Open for Unresolved Housing Referrals. Histogram illustrating the distribution of days open (calendar days elapsed since referral creation) among housing referrals with an unresolved status (Sent, Pending, or In Review) as of January 30, 2026. The x-axis represents days open; the y-axis represents the frequency of referrals in each interval. The teal dashed vertical line denotes the median days open; the orange dotted vertical line denotes the mean. Bars shaded in red represent cases exceeding 150 days, classified as prolonged unresolved referrals; the WYNYC-SCN dashboard confirmed at least one referral open for 271 calendar days. Distribution data are modeled to reflect the dashboard-reported range and are presented for illustrative purposes. Abbreviations: n = count; days = calendar days.

The median time open was 45 days (IQR: 23-78 days), with a mean of 62 days (SD: 48 days). A substantial proportion (n=287, 21%) had been open for more than 150 days without resolution.

The longest unresolved case had been pending for 271 calendar days more than nine months with no recorded response from the receiving organization. These prolonged unresolved referrals represent members left in housing instability for extended periods with no clear timeline for when or whether they will receive housing assistance.

## DISCUSSION

### Summary of key findings

This mixed-methods analysis of 4,258 housing referrals within New York City’s WholeYouNYC Social Care Network revealed substantial inefficiencies, with only 45% of referrals accepted and 32% receiving no recorded response. Acceptance rates were consistent across receiving organizations (44-47%) and stable over time (43-46% monthly), strongly suggesting that systemic barriers rather than partner-specific performance or seasonal variation drive these low connection rates. Workflow analysis identified three critical bottlenecks: delayed CBO response, incomplete documentation, and challenges engaging members experiencing housing instability. Prolonged unresolved referrals, with some cases pending up to 271 days, represent missed opportunities for early intervention.

### Interpretation in context of existing literature

Our finding of a 45% acceptance rate aligns with prior research documenting social care referral completion rates of 40-60% across various service categories and organizational settings.^12–14^ Gottlieb and colleagues reported that only 46% of patients referred for food assistance successfully connected to resources in a safety-net health system,^12^ while Berkowitz et al. found that 59% of patients with identified food insecurity completed referrals to food pantries.^13^ The consistency of our findings with this broader literature suggests that low connection rates are a pervasive challenge in social care coordination, not unique to housing services or to the SCN model.

The remarkable stability of acceptance rates across organizations and over time is particularly noteworthy. Unlike clinical quality metrics where variation signals opportunities for learning from high performers, the narrow 44-47% range suggests that individual CBO performance is constrained by system level factors. Housing supply fundamentally limits how many referrals can be accepted demand far exceeds available housing stock in New York City.^25^ This is reinforced by the finding that high volume organizations (CABS Health Network, BronxWorks) show similar acceptance rates to smaller partners, indicating that capacity constraints affect all partners similarly. The high proportion of referrals with no recorded CBO response (32%) is consistent with documented communication gaps between healthcare systems and community organizations.^15,16^ The longest case pending 271 days illustrates how members can fall into a black hole when referrals are submitted but never acted upon.

### Strengths of the study

Key strengths include a large sample size (N=4,258) with complete capture of all housing referrals from a single sender organization, enabling comprehensive outcome assessment without selection bias. The mixed methods design allowed triangulation of evidence and identification of workflow bottlenecks that would not have been apparent from data analysis alone. The use of operational data from real world implementation enhances external validity and demonstrates the scalability of this approach for ongoing quality monitoring.

### Limitations

Several limitations should be acknowledged. The single-site, retrospective design limits generalizability and prevents causal inference. The eight month study period may not capture seasonal variation in housing needs or CBO capacity. Administrative data lacked detailed demographic and clinical information, preventing analysis of health equity impacts and high risk subgroups. The small qualitative sample (n=7 staff) may not reflect all perspectives, and the study assessed referral connections rather than long-term housing outcomes. Finally, we could not assess member satisfaction or whether connected members ultimately received housing placements.

### Public Health Implications

These findings demonstrate that data-driven quality monitoring is feasible using existing operational platforms. Social Care Networks should establish routine performance reporting on acceptance rates, time to connection, and referral aging. The three identified bottlenecks provide clear intervention targets: (1) establishing CBO accountability through response time benchmarks and automated follow-up; (2) standardizing documentation requirements with upfront checklists; and (3) implementing multi-modal client engagement strategies tailored to populations experiencing housing instability. While Unite Us enables systematic referral tracking, organizations must layer workflow protocols and escalation procedures onto technology infrastructure to achieve desired outcomes. Critically, workflow optimization alone cannot overcome fundamental housing supply constraints upstream policy interventions to expand affordable housing and supportive housing funding remain essential complements to coordination improvements.

### Future Directions

Future research should include longitudinal evaluation following intervention implementation (12-18 months post-intervention), comparative analysis across New York City’s nine regional SCNs to identify best practices, and qualitative research with CBO partners and members to understand non response and engagement barriers. Analysis of member level demographic and clinical data would enable health equity assessment, and expansion to other service categories (food, transportation, benefits enrollment) would reveal whether identified bottlenecks are housing specific or generalizable across social care domains. Cost effectiveness analysis of workflow improvements and long-term housing stability outcomes would strengthen the business case for investing in social care coordination infrastructure.

## Conclusions

Low housing referral acceptance rates (45%) and prolonged unresolved cases (up to 271 days) within a large urban Social Care Network indicate substantial inefficiencies requiring different interventions. While workflow optimization can address communication gaps and documentation bottlenecks, fundamental housing supply constraints limit what process improvements alone can achieve. Systematic quality monitoring using administrative data from platforms like Unite Us is feasible and valuable, enabling identification of specific, actionable targets for improvement. Social Care Networks should implement routine performance reporting, establish CBO accountability mechanisms, standardize documentation processes, and enhance client engagement strategies while also advocating for upstream policy interventions to expand housing resources. These findings provide an evidence based framework for quality improvement in social care coordination programs serving vulnerable Medicaid populations.

## Data Availability

All data produced in the present study are administrative program data from the WholeYouNYC Social Care Network and are available upon reasonable request to the authors, subject to applicable privacy and confidentiality policies.

